# Impact of Executive and Adaptive function on functional outcomes in adults with Maple Syrup Urine Disease

**DOI:** 10.1101/2024.05.17.24307509

**Authors:** Jessica I Gold, Alanna Strong, Nina B Gold, Marc Yudkoff, Dava Szalda, Sophia Jan, Lisa A Schwartz, Rebecca Ganetzky

## Abstract

**Purpose:** Successful transition to adulthood requires intact executive and adaptive function. These neurocognitive domains, critical for independence, are frequently impaired in inherited metabolic disorders (IMD), though predictive clinical and biochemical factors are lacking. Standardized assessments linking neurocognitive status with functional outcomes are needed to improve prognostication and tailor support for affected emerging adults.

**Methods:** Maple Syrup Urine Disease (MSUD), a relatively prevalent IMD, is primarily diagnosed in the first week of life through newborn screening. Despite early intervention, executive and adaptive dysfunction persist. We designed a remote, interactive battery of neurocognitive and functional assessments for adults (≥21 years) with MSUD to correlate neurocognition and long-term outcomes.

**Results:** Assessments completed by 28 adults with MSUD (23 diagnosed after symptom onset) show a wide range in educational attainment, employment, and residence. Executive function, and adaptive function were significantly impaired in adults with MSUD compared to controls. Executive and adaptive deficits correlated negatively with obtaining skills needed for adult-oriented healthcare or independent living. Clinical history did not predict functional outcomes, but neurocognitive assessments suggest the benefits of pre-symptomatic diagnosis.

**Conclusion:** Independent adulthood is attainable for individuals with MSUD. Routine, targeted assessment of neurocognition may improve long-term functional outcomes in IMD.

## Introduction

Despite scientific and medical advances leading to improved health and cognition of people with inherited metabolic disorders (IMD), many affected individuals struggle moving from family-supported care to self-managed adult care.^1–4^ This transition requires executive function, which involves higher-level neurocognition like multi-tasking and behavior regulation, and adaptive function, which manages daily living demands.^5^ These skills are critical for independent adulthood, especially for those with complex medical diagnoses.^6–10^ Young adults with deficits in these neurocognitive domains often experience lower treatment adherence, school readiness, and employment retention.^6,11^ However, little is known about how these neurocognitive processes specifically impact functional outcomes in the IMD population, a factor that limits the efficacy of evidence-based interventions to improve independence and autonomy.

For many with IMD, executive and adaptive dysfunction persists despite early identification and treatment. Early treatment of phenylketonuria, for example, prevents intellectual disability, yet adults continue to demonstrate executive dysfunction that does not reliably correlate with phenylalanine level.^12,13^ Nearly 30% of asymptomatic woman with ornithine transcarbamylase deficiency show deficits in working memory.^14^ Similarly, liver transplantation in urea cycle disorders prevents episodic hyperammonemia but has not consistently improved long-term cognitive outcomes.^15^ These challenges highlight the complexity of linking biomarkers, clinical history, genotype, and long-term functional outcomes, complicating our ability to provide anticipatory guidance.

To address these gaps, we examined the impact of executive and adaptive dysfunction on functional outcomes in adults with Maple Syrup Urine Disease (MSUD), a relatively prevalent IMD with an easily identifiable biochemical signature that allows for early diagnosis and intervention, leading to improved health outcomes.^16^ Deficiency of branched-chain amino acid metabolism leads to accumulation of disease-specific biomarkers (valine, isoleucine, leucine, and alloisoleucine), allowing for pre-symptomatic diagnosis through biochemical newborn screening.^16,17^ Elevated branched-chain amino acids can cause acute intoxication with encephalopathy and life-threatening cerebral edema, as well as a chronic, neurodegenerative phenotype with executive dysfunction, intellectual impairment, and mood and attention disorders.^16,18^ Careful control of amino acids via diet, treatment with the co-factor thiamine and/or prompt medical intervention during periods of illness can minimize hyperleucinosis and improve long-term outcome.^17^ Liver transplantation can restore peripheral BCAA metabolism and stabilize neurocognitive function, but may not prevent neuropsychiatric complications.^16,19,20^

Previous studies suggest underlying executive and adaptive dysfunction in MSUD patients (with younger patients showing higher IQ potentially due to pre-symptomatic diagnosis.^21^ However, discrepancies between verbal and functional IQ persistent, a pattern that is indicative of executive dysfunction.^18,22^ Liver transplantation has been shown to improve IQ score but not adaptive functioning.^19^ Adolescents with MSUD struggle with activities requiring intact executive adaptive function, indicating challenges in transitioning to independence.^23^

Neurocognitive and psychosocial outcomes in MSUD have not strongly correlated with biochemical or clinical history.^16^ We hypothesize that standardized neurocognitive assessment will improve the prediction of functional outcome, including which MSUD patients can most successfully manage their underlying disease. Therefore, using a novel remote study design, we assessed the functional outcomes in a cohort of adults with MSUD, including educational attainment, employment, and living situation, and correlated outcomes with a neurocognitive battery quantifying executive and adaptive dysfunction. We used subgroup analysis to examine the effects of pre-symptomatic diagnosis and liver transplantation. Further, we enrolled a racially and ethnically diverse cohort to improve generalizability over previous studies that focused on homogenous populations. Together, this study addresses knowledge gaps on the long-term outcomes for adults with MSUD, providing better guidance on therapeutic supports and enhancing our ability to predict disease prognosis.

## Material and Methods

### Participants

This study was approved by the Institutional Review Board at the Children’s Hospital of Philadelphia (CHOP) (IRB#21-18433). Eligible participants with MSUD were aged 21 years or older. Fluency in English was an admission requirement in order to conform to the criteria of the cognitive measures. Exclusion criteria were moderate to severe intellectual disability. Control participants were recruited via two methods. Each adult with MSUD was asked to identify one unaffected acquaintance or relative of a similar age to serve as a paired control. For participants who were unable to provide contact information, recruitment of controls was aided by the Recruitment and Cohort Selection (REC) Sub-Core of the Intellectual and Developmental Disabilities Research Center at CHOP. A recruitment email was sent to eligible participants in the REC database of healthy volunteers. Interested individuals contacted the study team by email or phone and completed the same consent process. All participants completed the same battery of neurocognitive and psychosocial outcomes assessments.

### Procedures

Potential participants were informed about the study through the MSUD Family Support Group newsletter, postings on social media, or an information session organized by the young adult members of the MSUD Family Support Group. Informed consent was obtained via electronic signature of a written consent form via Redcap.^24^ Participants received 3 reminders by email or text to complete study outcomes measures. Consort flow diagram for eligibility and recruitment are in Fig S1.

### Metabolic history

Medical records were obtained for individuals with MSUD. Information excerpted from medical history included: method and age of diagnosis, presence of symptoms at diagnosis, genotype, frequency of hospital admissions, method of MSUD treatment, history of liver transplant, and presence of comorbid affective health disorders. The frequency of plasma amino acid measurements varied among participants, therefore calculations of median blood leucine concentration for each 12-month period of life was utilized, as has been previously used for other IMD that involve nutritional toxicity.^12,25^

### Measures

All measures (Table S1) were selected based on their ability to be completed remotely due travel limitations during the COVID-19 pandemic. For questionnaires with multiple reporters, participants were asked to designate an informant with whom they regularly interacted in the last month, including parents, caregivers, siblings, or roommates. The BRIEF-A and ABAS-3 surveys were administered and interpreted by the CHOP Center for Human Phenomic Science Behavioral Core. The Adaptive Cognitive Evaluation (ACE) is a web-based platform that uses adaptive games to personalize the assessment for each participant’s abilities.^26^ It has previously been used to assess executive function skills in individuals with multiple sclerosis, dementia, 16p11.2 deletion syndrome, and sensory processing disorders.^26–29^ Participants were able to access the ACE remotely using a web browser or through apps designed for Apple or Android phones.^30^

### Data Handling

ACE Data Cleaning: Data from each participant were evaluated and cleaned using the dedicated R package Neuroscape’s aceR analytics package as previously described.^30^ Filtering data on a game-level basis prevents wholescale exclusion of participants, while including only data for which the participant understood and complied with game instructions.

### Statistical Analysis

Data were summarized using descriptive statistics (mean and standard deviation for continuous variables and proportion for categorical variables). Categorical variables were analyzed using Chi-square (or Fisher’s when appropriate), T-tests, or analysis of variance (ANOVA). Linear regression modeling of continuous variables (survey scores, ACE game scores, leucine levels) was performed adjusting for age. For all analyses, two-sided p-values <0.05 that surpassed the Bonferroni-corrected significance threshold for multiple testing were considered significant and noted with an asterisk. All analyses were performed with R software, version 4.2.2. (R Project for Statistical Computing).

## Results

### Participants

Demographic information for affected and control participants are found in Table 1. There was no significant difference in age (t=0.098, p=0.92), gender (x^2=2.34, p=0.13), years of education (t=1.5, p=0.14), or income (t=0.68, p=0.51) between the two groups. Participants in the control group were more likely to be employed full-time (x^2=4.92, p=0.03, more likely to live with a roommate (x^2= 5.1, p<0.02), and less likely to live with their parents (x^2= 9.39, p<0.002). Both groups endorsed similar access to medication and transportation to medical appointments.

**Table 1:** Demographics.

|  | MSUD | Control |
| --- | --- | --- |
| Gender, n (%) |  |  |
| Male | 10 (35.7) | 7 (31.8) |
| Female | 18 (64.3) | 15 (68.2) |
| Age, mean (SD) | 33.4 (7.1) | 33.6 (7.1) |
| Race/Ethnicity, n (%) |  |  |
| Asian | 1 (3.5) | 1 (4.5) |
| Black | 2 (7.2) | 4 (18.2) |
| White/Hispanic | 4 (14.3) | 1 (4.5) |
| White/non-Hispanic | 21 (75) | 16 (72.3) |
| Residence, n (%) |  |  |
| Alone | 5 (17.9) | 5 (22.7) |
| Domestic Partnership | 10 (35.7) | 10 (45.5) |
| Parents | 12 (42.9)* | 1 (4.5)* |
| Roommate | 1 (3.5)** | 6 (27.3)** |
| Marital Status, n(%) |  |  |
| Single | 15 (53.6) | 10 (45.5) |
| Cohabitation | 2 (7.1) | 0 (0) |
| Married | 9 (32.1) | 10 (45.5) |
| Divorced | 2 (7.1) | 2 (9.1) |
| Children, n (%) | 7 (25) | 9 (40.9) |
| Mean years education (SD) | 15.3 (2.8) | 16.3 (1.73) |
| Employment, n (%) |  |  |
| Full-time employment | 9 (32.1)<sup>†</sup> | 14 (63.6)<sup>†</sup> |
| Part-time employment | 8 (28.5) | 4 (18.2) |
| Unemployed | 9 (32.1) | 2 (9.1) |
| Student | 2 (3.6) | 2 (9.1) |
| Income, n(%) |  |  |
| Below \$10,000 | 6 (21.4) | 0 (0) |
| \$10,000-50,000 | 1 (3.6) | 5 (22.7) |
| \$50,000-100,000 | 5 (17.9) | 7 (31.8) |
| \$100,000-150,000 | 5 (17.9) | 4 (18.2) |
| Over \$150,000 | 2 (7.1) | 5 (22.7) |
| Prefer not to say | 5 (17.9) | 0 (0) |
| I don't know | 4 (14.3) | 1 (4.5) |
| Medication Insecurity Frequency, n (%) |  |  |
| 0 times in past year | 17 (60.7) | 15 (68.2) |
| 1-5 times in past year | 8 (28.6) | 4 (18.2) |
| 6-10 times in past year | 2 (7.1) | 2 (9.1) |
| >10 times in past year | 1 (3.6) | 0 (0) |
| Formula Insecurity Frequency, n (%) |  |  |
| 0 times in past year | 17 (60.7) | - |
| 1-5 times in past year | 9 (32.1) | - |
| 6-10 times in past year | 1 (3.6) | - |
| >10 times in past year | 1 (3.6) | - |
| Lack of Transportation for Appointments Frequency, n (%) |  |  |
| 0 times in past year | 23 (82.1) | 19 (86.4) |
| 1-5 times in past year | 2 (7.1) | 2 (9.1) |
| 6-10 times in past year | 3 (10.7) | 1 (4.5) |
| >10 times in past year | 0 (0) | 0 (0) |
| Liver Transplant | 5 (17.9) | - |
| Symptomatic at Diagnosis | 23 (82.1) | - |

Chi-squared comparisons showed significant difference between affected and control participants: \* $\chi^2= 5.1$ , $p<0.02$ ; \*\* $\chi^2= 9.39$ , $p<0.002$ ; † $\chi^2=4.92$ , $p=0.03$

### Functional Outcomes

Affected participants were divided into three groups: (1) symptomatic diagnosis with native liver (SD), (2) symptomatic diagnosis status post liver transplantation (SD/LT), and (3) diagnosis prior to symptom appearance (asymptomatic diagnosis, AD)^18^. The symptomatic diagnosis group consisted of 23 individuals (82.1%) who initially presented with symptoms ranging from neonatal encephalopathy to failure to thrive. The asymptomatic diagnosis group (n=5, 17.9%) included one individual diagnosed via newborn screening and four individuals, three with classical MSUD and one with late-onset MSUD, who were identified pre-symptomatically following sibling diagnosis. 17.9% of participants (5/28), all diagnosed symptomatically, were recipients of liver transplants with an average age of transplantation at 17.6±9.69 years.

Social outcomes for affected participants are reported in Fig 1A-D. 25% completed a graduate degree (7/28); 6 with master’s degrees, 1 with a PhD), 28.5% (8/28) graduated from college, 25% (7/28) started college but did not graduate, 14% (4/28) had a terminal high school degree, and 7.1% with some high school completed (2/28) (Fig 1A). Employment status was evenly distributed with 32.1% employed full-time (40+ hours per week) (9/29), 28.5% are employed part-time (less than 40 hours weekly) (8/28), and 32.1% are unemployed (9/28) with 2 individuals (7.1%) indicating they are full-time students (Fig 1B). Most participants (53.5%, 15/28) had never been married while 39.2% (11/28) were currently either married or cohabitating with a romantic partner (Fig 1C). 42.9% (12/28) of participants resided with their parents while 35.7% (10/28) lived with a domestic partner, 17.9% (5/28) lived alone and 3.6% (1/28) lived with a roommate (Fig 1D). Participants reported a wide range of annual incomes (Fig S2). For all social outcomes, there were no significant differences between the three clinical history-based subgroups (symptomatic diagnosis with native liver, symptomatic diagnosis post-transplantation, and asymptomatic diagnosis). For the sixteen patients with available leucine levels, there was no correlation between overall mean leucine, mean childhood leucine (levels from ages younger than 18 years), mean adult leucine (levels from age 18 years and older), or peak reported leucine on any functional outcome (Table S2).

**Fig 1.**
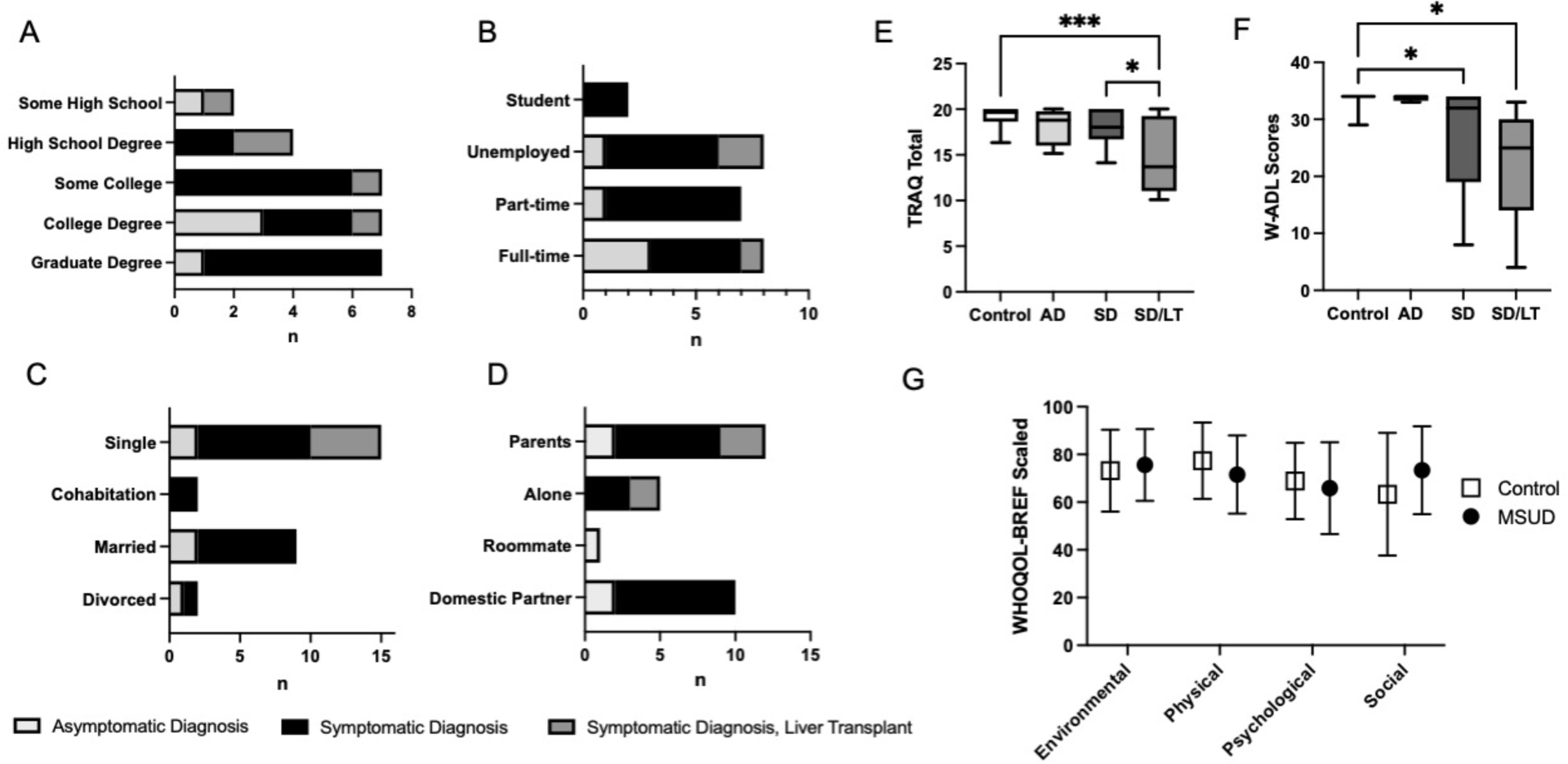
Surveying psychosocial outcomes for adults with MSUD show wide variability without correlation to medical history, while skills for independent living and medical self-management do relate to medical history. Participant responses to a demographic survey were classified by method of MSUD diagnosis and history of liver transplantation. Participants reported current marital status (A), residence (B), terminal degree (C), employment (D), and income (E). Full-time employment was defined as equal to or greater than 40 hours per week. Part-time employment was defined as less than 40 hours per week. (F) Total scores for self-report of medical self-management via the TRAQ showed a significantly lower scores in affected individuals diagnosed symptomatically compared to controls (\**p=0.027*), and liver transplant recipients compared to controls (\*\*\**p=0.0002*). (G) Scaled scores for self-report of the W-ADL showed a significant difference between the control cohort and individuals with symptomatic diagnosis and native livers (\**p=0.033*) and the control cohort and individuals with symptomatic diagnosis and transplanted livers (\**p=0.024*). (H) Scaled scores for the WHOQOL-BREF survey show no difference in any quality of life domain between the control population and participants with MSUD. *SD/NL: Symptomatic Diagnosis, native liver; SD/LT: Symptomatic Diagnosis/Liver Transplant*

Participants were surveyed about the skills for activities of daily living (ADL) via the W-ADL and medical self-management using the Transition Readiness Assessment Questionnaire (TRAQ).^31,32^ Adults with symptomatic diagnoses of MSUD had significantly lower scores on the W-ADL, but not the TRAQ (Fig 1E-F), indicating fewer ADL-related skills, but sufficient skills for medical self-management. Liver transplantation correlated with significantly lower scores in both questionnaires compared to controls and on the TRAQ compared to the symptomatic cohort with their native liver. Older age was positively correlated with total TRAQ score (Table S1) but did not correlate with W-ADL score. Increased educational attainment also correlated with overall scores on the TRAQ and the W-ADL plus scores on TRAQ subdomains for medication management, appointment keeping, and tracking health issues (Table S1).

Quality of life was surveyed using the WHOQOL-BREF. There was no difference between our control group and our affected group in any of the measured quality of life domains (Fig 1G). There was no correlation between reported quality of life and method of diagnosis, liver transplant, or age, but there was a positive correlation between educational attainment and the WHOQOL social subdomain (Table S1). As above, leucine levels did not correlate with any measured outcome (Table S3).

### Executive Function Outcomes – BRIEF-A

Compared to control, adults with MSUD had significantly higher mean T-scores (signifying worse executive function) in the General Executive Composite (GEC, Fig 2A, t=4.26, p=0.0001) and the 2 sub-indices: Metacognition index (MI, Fig S3A t=5.19, p<0.001), and the Behavioral Regulatory Index (BRI, Fig S3B, t=3.11, p=0.003). GEC T-scores for participants with MSUD remained just inside the normal range (58.9±13.6), while mean T-scores for MI (62.9±13) and GEC (61.3±13.3) were in the mildly elevated range. BRIEF-A subscales showed 5 domains with T-scores in the mildly elevated range (Fig S3C).

**Fig 2.**
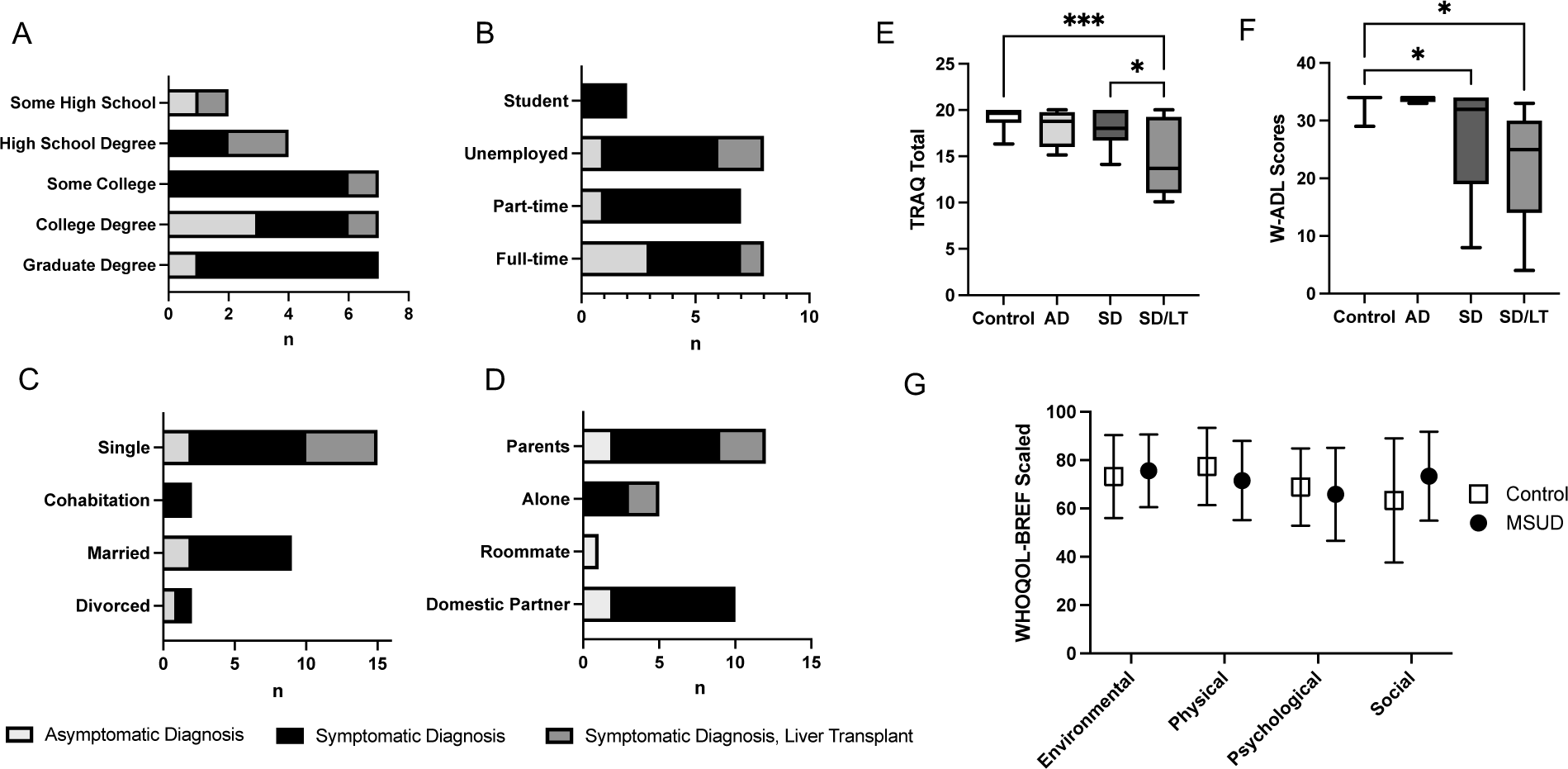
The BRIEF-A questionnaire showed greater deficits in executive function in adults with MSUD and correlation between age-normalized scores and functional outcomes. (A) Adults with MSUD had significantly higher General Executive Composite (GEC) composite T-scores, signifying greater executive function deficits (\*\*\**p=0.0001*). (B) GEC T-scores did not correlate with current residence for participants with MSUD. (C) For affected participants, GEC T-score was significantly higher, signifying more executive dysfunction, for individuals with a terminal high school degree than those who completed a graduated degree (*\*\*p=0.0049, *p=0.016*). (D) For affected participants, GEC T-score significantly lower, signifying better executive function for those working full-time (40 hours per week and greater) versus those who are students (\*\**p=0.*0049), those unemployed (\*\**p=0.*0012), and those working part-time (less than 40 hours per week, \*\*\**p=0.0005*). Forest plots are shown illustrating the regression slope with 95% confidence intervals and p-values for regression models adjusted for age between BRIEF-A T-scores and total TRAQ scores (E) and scaled W-ADL scores (F). All p-values noted with an asterisk are significant beyond the Bonferroni-adjusted scale of 0.025. (G) Analysis of GEC T-scores based on diagnosis method showed a significantly higher scores, signifying more executive dysfunction for individuals diagnosed symptomatically (SD) with MSUD compared to controls (\*\*\**p=0.0002*). (H) There is no significant different in GEC T-scores for individuals who receives a liver transplant (LT) compared to those with their native liver (NL) and both are significantly higher (worse executive function) than controls. (**p<0.01) *GEC: General Executive Composite; AD: Asymptomatic Diagnosis; SD: Symptomatic Diagnosis; NL: Native Liver; LT: Liver Transplant*

ANOVA and regression modeling was used to determine the relationship between executive dysfunction and functional outcomes. GEC T-score correlated with completing a graduate degree (Fig 2B) and being employed full-time (Fig 2C). It did not correlate with place of residence (Fig 2D). Linear regression modeling, corrected for age, showed a correlation between lower T-scores (signifying better executive function) and more years of education completed (Fig S3D). There was also a modest correlation between TRAQ scores and all BRIEF-A indices (Fig 2E), and W-ADL scores and the 2 subindices (MI, BRI), but not the main composite score (Fig 2F), signifying a negative relationship between executive dysfunction and skills required for medical and functional autonomy. Executive function had no significant impact on quality of life for adults with MSUD. There was only a significant correlation between the Organization of Materials subscale and median leucine levels in adulthood (Table S3).

Our data suggests that asymptomatic diagnosis leads to improved T-scores on all BRIEF-A scales as significant difference exists between adults diagnosed symptomatically with MSUD and our control group which is not present in our group diagnosed asymptomatically (Fig 2G). Liver transplantation does not improve scores on the BRIEF-A compared to controls (Fig 2H).

### Executive Function Outcomes – Games

#### Attention

Adults with MSUD scored significantly lower, indicating worse general attention skills, than the control participants in Face Switch (Fig 3A, t=2.279, p=0.029), Boxed (Fig 3B, t=3.35, p=0.0018), and Flanker (Fig 3C, t=3.2, p=0.0029). There was no significant difference in mean reaction time (MRT) between affected participants and the control group when assessing sustained attention (Fig 3D, t=1.93, p=0.061) or impulsive attention (Fig 3E, t=1.66, p=0.11). *Working Memory:* Adults with MSUD scored significantly worse than controls in all 5 games: Spatial Cue (Fig 3F, t=2.28, p=0.029), Stroop (Fig 3G, t=3.42, p=0.0016), Forward span (Fig 3H, t=3.88, p=0.0004), Backward span (Fig 3I, t=2.15, p=0.038), and Filter (Fig 3J, t=2.19, p=0.035). *Goal Management:* Adults with MSUD scored significantly worse compared to control in Triangle Trace (Fig 3K, t=2.73, p=0.01) and Task Switch (Fig 3L, t=2.53, p=0.016).

**Fig 3.**
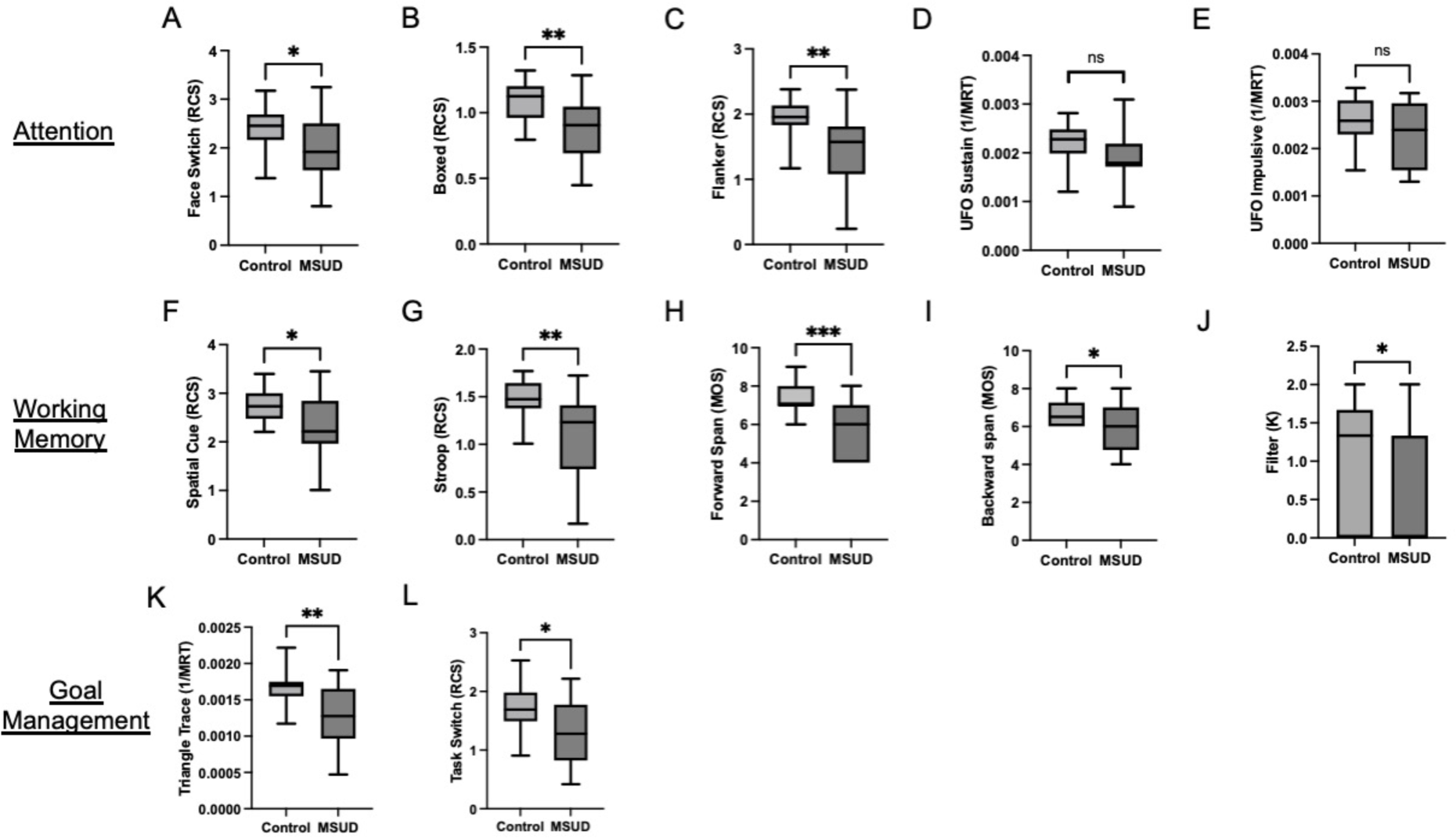
The novel, virtual platform of the Adaptive Cognitive Exam (ACE) demonstrated executive function deficits for adults with MSUD across all three subdomains. Data for the 12 ACE games was analyzed using the aceR package. The designated metric for each game is presented. Executive function deficits are indicated by lower Rate Correct Score (RCS), lower inverted Mean Reaction Time (1/MRT) or lower Mean Object Span (MOS) indicates executive function deficits. K is a measure of working memory capacity where higher values demonstrate better capacity. above. \**p<0.01; **p<0.005; ***p<0.0005* *RCS: Rate Correct Score; MRT: Mean Reaction Time; MOS: Maximum Object Span*

Correlation between each ACE game and functional outcomes was performed. From this analysis, we identified ACE games that correlated with functional status: residence (Face Switch, Impulsive Attention, Stroop, Spatial Cue, Fig S4A); terminal degree (Boxed, Sustained Attention, Impulse Attention, Stroop, Fig S4B); employment (Face Switch, Boxed, Impulsive Attention, Task Switch, Backward span, Stroop, Spatial Cue, Fig S4C). Regression modeling controlled for age showed positive correlation between total TRAQ score and scores on Face Switch, Boxed, Impulsive Attention, Sustained Attention, Triangle Trace, Stroop, Task Switch, Forward Span and Backward Span (Fig S4D). W-ADL scores correlated positively with scores in Boxed and Face Switch (Fig S4E). Historical leucine levels did not correlate with scores in any ACE game (Table S5). Presence of symptoms at MSUD diagnosis did not impact scores on any of the ACE games. However, on six games (Face Switch, Boxed, Stroop, Task Switch, Forward Span, and Backward span), there was a significant difference in score between individuals diagnosed symptomatically and the control group that was not present in those diagnosed asymptomatically (Fig S4F). Individuals who received liver transplants scored significantly worse in five games (Boxed, UFO Sustained, UFO Impulsive, Triangle Trace, and Forward Span) compared to individuals with their native livers (Fig S5).

### Adaptive Function Outcomes – ABAS-3

Adaptive function was assessed using the ABAS-3. Compared to controls, affected participants scored significantly lower signifying worse adaptive function on the composite and all three subdomains: GAC (t=2.94, p=0.0052), Conceptual (t=3.07, p=0.0038), Practical (t=3.83, p=0.004), and Social (t=3.96, p=0.0003). (Fig 4A, S6A-C); however, the mean scaled scores for each component remained in the lower end of the normal range (Conceptual: m=93±17.4; Practical m=92.3±17.5: Social m=92.8±15.9; GAC m=95.3±18.4). Individuals living with spouses scored significantly higher in the GAC (better adaptive function, Fig 4B). Individuals with terminal college or graduate school degrees scored significantly higher (better adaptive function) in the GAC compared to those with terminal high school degrees (Fig 4C). Adults employed full-time also scored significantly higher (better adaptive function) on all scales compared to those employed part-time (GAC, Fig 4D). Linear regression modeling corrected for age showed positive correlation between all 4 scales and total TRAQ score (Fig 4E), and between GAC, conceptual, and practical scores and total WADL score (Fig 4F). There was no correlation between leucine levels and any ABAS-3 scale (Table S6). GAC scores were significantly lower (worse adaptive function) for adults diagnosed after symptom onset compared to control group while no statistical difference was present in the asymptomatic diagnosis cohort (Fig 4G). History of liver transplantation did not improve ABAS-3 scores compared to the native liver cohort (Fig 4H).

**Fig 4:**
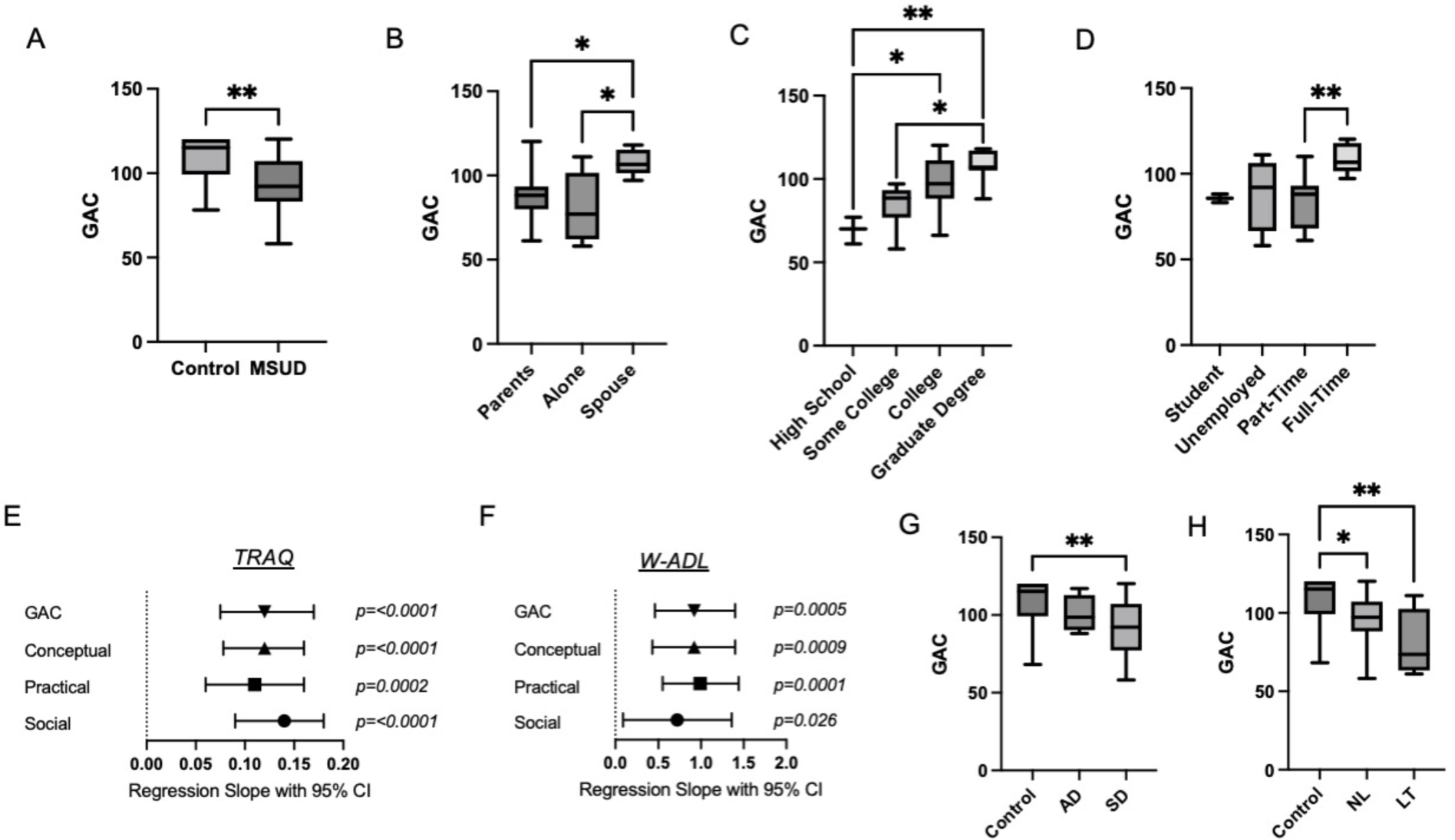
Assessment of adaptive function using the ABAS-3 questionnaire showed adaptive function deficits in adults with MSUD that correlate with functional outcomes and skills for independent living. (A) Adults with MSUD had a significantly lower General Adaptive Composite (GAC) compared to the control group, signifying greater deficits in adaptive function. \*\**p<0.001* (B) Adults living with a spouse had significantly higher GAC (better adaptive function) compared to those living with parents or living alone. \**p<0.01* (C) Terminal degree correlated with GAC score. Individuals with only a high school education had significantly lower GAC (worse adaptive function) than those with terminal college or graduate degrees. Adults who attended some college had a significantly lower GAC (worse adaptive function) than those with graduate degrees. \**p<0.01; *p<0.001* (D) Adults employed full-time had significantly higher GAC (better adaptive function) compared to those employed part-time. \*\**p<0.001* (E-F) Forest plots are shown illustrating the regression slope with 95% confidence intervals and p-values for regression models adjusted for age between GAC score and total TRAQ scores (E) and scaled W-ADL scores (F) Both plots show a relationship between better adaptive function and higher scores on skills assessments. All p-values noted with an asterisk are significant beyond the Bonferroni-adjusted scale of 0.025. (G) Adults who were diagnosed with MSUD symptomatically (SD) had significant lower GAC (worse adaptive function) compared to the control group. There was no significant difference in GAC between the control group and individuals diagnosed prior to symptom appearance (AD). \*\**p<0.001I* (H) Liver transplantation did not improve GAC compared to control as both individuals with their native livers (NL) and transplant recipients had significantly lower GAC (worse adaptive function). \**p<0.01, **p<0.001* *AD: Asymptomatic Diagnosis; SD: Symptomatic Diagnosis; NL: Native Liver; LT: Liver Transplant*

## Discussion

This is the first study to systematically correlate neurocognitive function with real-life outcomes for adults with MSUD, using a complementary set of validated interactive assessments. Our findings show a wide variation in the educational attainment, employment, and living situation for adults with MSUD. Participants with MSUD demonstrated executive dysfunction with specific deficiencies in metacognition domains. Adaptive function assessment revealed mean scores at the lower end of normal range, but significantly lower than age-matched, neurotypical siblings and peers. Importantly, individuals with better executive or adaptive function were more likely to score highly on questionnaires on skills needed for adult-oriented healthcare or independent living. Clinical history, including timing of diagnosis, history of liver transplant, or median leucine levels, did not predict functional outcomes. However, our neurocognitive assessments suggested a benefit for diagnosis and treatment of MSUD prior to symptom appearance. Overall, these findings expand our understanding of the long-term functional outcomes of people with MSUD and exemplify the need to follow adult outcomes in all IMD.

### Why focus on executive and adaptive function?

Executive and adaptive function are being incorporated in healthcare transition readiness assessments for other complex conditions of childhood such as sickle cell disease and spina bifida.^33,34^ Both are modifiable processes that impact long-term functional outcomes independent of intellectual ability.^6,35,36^ Integrating executive function assessment and training into healthcare transition (HCT) planning may ease the path to independence. Our study suggests an importance for metabolic clinicians in HCT: none of the transplanted participants was followed at a metabolic clinic post-procedure; this cohort showed significantly lower TRAQ scores than the symptomatic diagnosis/native liver cohort. However, as current HCT planning for IMD primarily emphasizes dietary management, a more holistic, inclusive approach to transition readiness is needed.^37^ Metabolic clinicians should lead in designing and implementing HCT interventions that focus on skills for medical self-management and independent living as well as the neurocognitive capabilities to obtain these skills.

### Identifying Prognostic Factors

Our results suggest that the presence of symptoms at MSUD diagnosis significantly predicts greater executive and adaptive dysfunction, but no significant difference in functional outcomes. This result may be confounded by the finding that 80% of participants with asymptomatic diagnoses had a previously affected sibling. It is possible that these families applied previously acquired knowledge on MSUD management, which may have improved their child’s health in early infancy and childhood. Analysis of the impact of clinical history was limited by challenges in obtaining lifelong medical records. In many instances, our patients’ date of birth preceded widespread adoption of electronic health records, thus comprehensive documentation of their early health history was infrequent. Only 9 (32.1%) participants had documented genotypes, constraining any genotype-phenotype correlations. In future work, we will expand this protocol to younger participants, where greater availability of lifelong detailed medical records may aid in identifying prognostic factors.

### Impact of Liver Transplantation

Our transplanted participants had significant executive and adaptive dysfunction, despite 4 (80%) undergoing transplantation in early adolescence, prior to the acquisition of many executive and adaptive function skills.^5,38^ Based on interviews with patients and their families, liver transplantation tended to occur only after patients demonstrated persistent poor dietary compliance, frequent decompensation, or strong parental advocacy. Thus, transplanted individuals may have had previous neural injury from hyperleucinosis that prevented maturation of executive and adaptive function skills. Further, recent studies show that extrahepatic MSUD manifestations, including effects on neurotransmitter biochemistry, may not be ameliorated by liver transplantation.^16^ Our results support maintaining care continuity post-transplant with metabolism providers. These individuals would benefit from a less diet-centered approach to MSUD care that emphasizes monitoring of neuropsychiatric sequelae and developing skills for medical self-management and independent living.

### Limitations

The main limitation of this study was generalizability of results. Our recruitment strategy required some modicum of executive function for enrollment. Affected individuals were required to initiate contact with the research team, receive all communication, and complete the questionnaires and ACE. Thus, it is possible that we recruited individuals who had relatively intact preservation of executive function. While the proportions may be skewed toward our higher-functioning participants, the data presented here shows persistent executive and adaptive dysfunction that impact functional outcomes. Notably, our cohort did not include any North American Old Order Mennonite participants. The functional outcomes assessed here may not be representative of the unique structure of long-term care in these communities.

### Future Directions

Expansion of this study to younger individuals with MSUD, specifically those diagnosed by newborn screening and those who received liver transplantation in early childhood, will further define prognostic factors. Future studies will utilize a MSUD-specific subset of ACE games that are highly correlated with functional outcome. This MSUD-specific ACE will be used contemporaneously with branched-chain amino acid analysis to examine the influence of leucine level on game scores - perhaps, providing a proxy for hyperleucinosis that requires medical intervention. We will also explore applicability of our novel neurocognitive battery in other IMD that lack determinants of functional outcomes in adulthood.

## Conclusion

Advances in the diagnosis and treatment of IMD have fundamentally altered the health of people with these disorders. Using a novel remote study design, we assessed neurocognitive and functional outcomes for a diverse cohort of adults with MSUD. Our data showed significant executive and adaptive dysfunction in adults with MSUD compared to controls. Neurocognitive function correlated with social outcomes, including education and employment. Overall, this study demonstrates that milestones of adulthood are attainable for individuals with MSUD, though better support for executive and adaptive function are needed. This study provides a model for describing the impact of neurocognitive status on functional outcomes and may be highly applicable across rare diseases.

## Data Availability

All data produced in the present study are available upon reasonable request to the authors

