## supplemental figures for "Impact of Executive and Adaptive function on functional outcomes in adults with Maple Syrup Urine Disease"

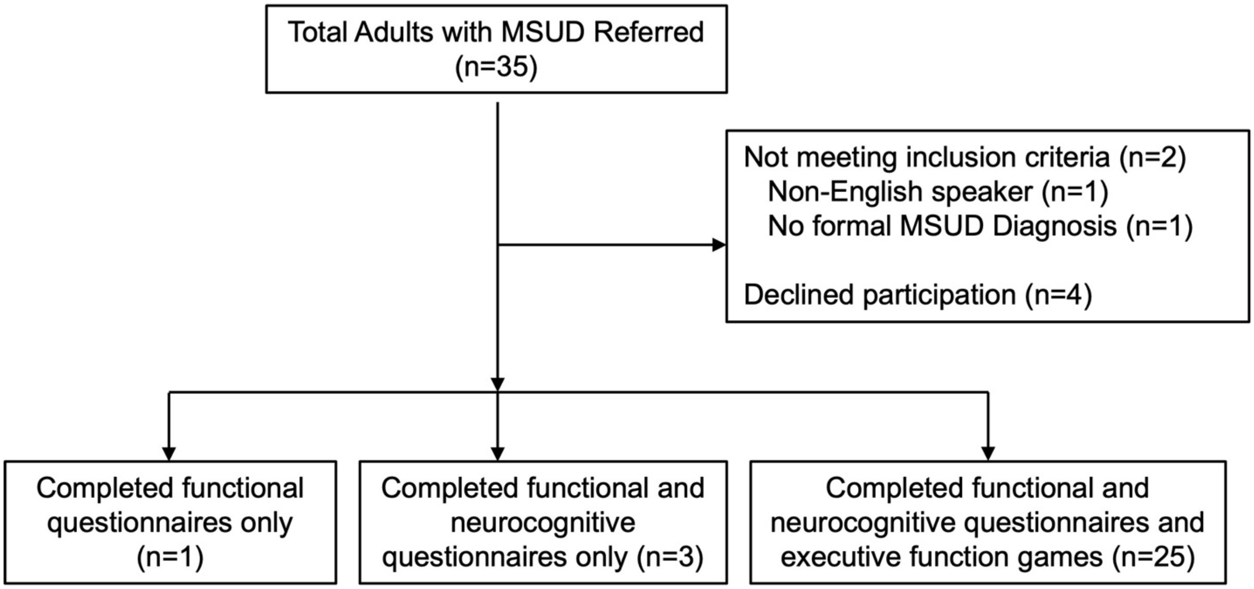


Fig S1: Consort flow diagram of recruitment and enrollment.

Table S1: Neurocognitive and Psychosocial Measures

| Measure | Format | Items | Measured Domains | Scoring | Interpretation | References |
|---|---|---|---|---|---|---|
| Behavior Rating Inventory of Executive Function for Adults (BRIEF-A) | Electronic Questionnaire with self-report and informant-report forms | 75 | General Executive Composite (GEC); Behavioral Regulation Index (BRI); Metacognition Index (MI); 9 subscales: Inhibit, Self-Monitor, Plan/Organize, Shift, Initiate, Task Monitor, Emotional Control, Working Memory, and Organization of Materials | Age-normalized T-score with mean 50 and standard deviation; $T \leq 59$: normal T = 60-64: Mildly elevated $T \geq 65$: Significantly Elevated | Higher scores correlate with greater executive function deficits | |
| Adaptive Behavior Assessment System, Third Edition (ABAS-3) | Electronic Questionnaire with self-report and informant-report forms | 239 | General Adaptive Composition (GAC); Conceptual Domain; Social Domain; Practical Domain | Age-normalized scaled scores range from 40-130 Scores < 90: Below average Scores 90-110: Average Score >110: Above Average | Lower scores correlate with greater adaptive function deficits | |
| Adaptive Cognitive Evaluation | Web-based adaptive games | 12 games lasting 2-5 minutes each | Response Time; Working Memory; Controlled Attention; Context Monitoring; Interference Resolution; Cognitive Flexibility | Detailed game analysis and scoring can be found in Younger et al. | Rate Correct Score (RCS), Mean Object Span (MOS), K: lower scores correlate with greater executive function deficits Mean Response Time (MRT): higher scores correlate with greater executive function deficits | |
| Waisman Activities of Daily Living (W-ADL) | Electronic Questionnaire with self-report and informant-report forms | 17 | Uses 3-point scale to rate current level of independence in: Grooming, Toileting, Feeding, Meal Preparation, Household chores, Errands, Household repairs | Scores on each item range from "0 - does not do at all," "1– does with help," "2 – Independent or does on own" Scores range from 0-34 | Higher scores indicate greater ability to complete activities of daily living | |
| Transition Readiness Assessment Questionnaire (TRAQ) | Electronic Questionnaire with self-report and informant-report forms | 20 | Uses 5-point scale to evaluate skills for managing medications, appointment keeping, tracking health issues, and talking with providers | Composite scores range from 5 – "I do not know how" to 20 – "Yes, I always do this when I need to" | Higher scores correlate with greater attainment of skills needed for medical self-management | |
| WHOQOL-BREF | Questionnaire with self-report and informant-report forms | 26 | Uses 5-point scale to evaluate physical health, psychological health, social relationships, environment | Scores linearly transformed to 0-100 point scale | Higher scores correlate with higher quality of life | |

Figure S2

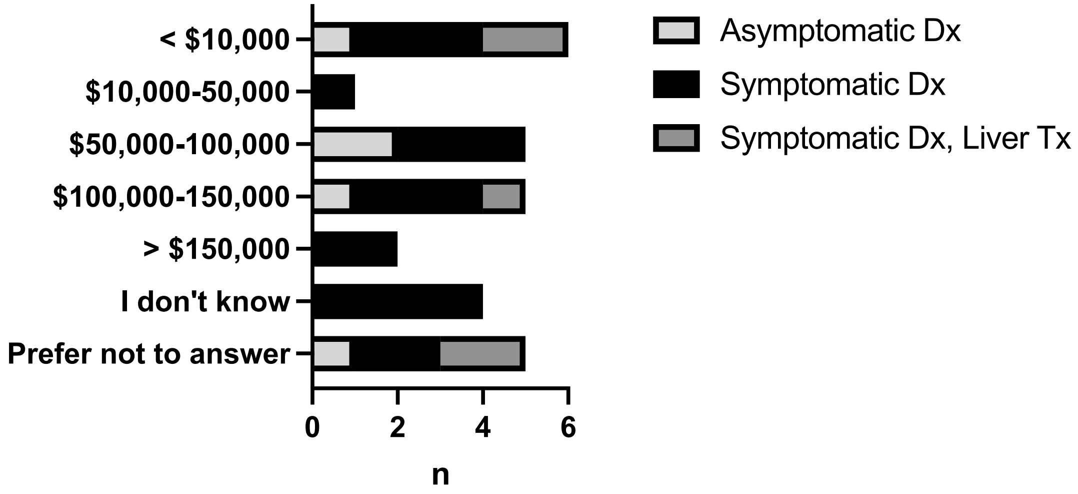


Fig S2: Adults with MSUD report a wide range of annual incomes that do no correlate with clinical history.

Table S2: Historical Leucine Levels and Functional Outcomes

| | Mean Lifetime Leucine (SD) | Mean Childhood Leucine (SD) | Mean Adult Leucine (SD) | Peak Leucine | n |
|---|---|---|---|---|---|
| **Education** | | | | | |
| HS Degree | 373.4 (200.5) | 325 (281.5) | 352.1 (172.9) | 838.3 (437.7) | 5 |
| Some College | 355 (30.2) | 414 (217.3) | 343.2 (13.7) | 667.4 (279.5) | 2 |
| College Degree | 183.2 (60.1) | 225.6 (235.8) | 193.2 (68.6) | 934.5 (102.5) | 2 |
| Graduate Degree | 385.2 (208.4) | 336.8 (213.8) | 383.9 (210) | 753.6 (330.5) | 5 |
| **Employment** | | | | | |
| Full-time | 434.7 (222.4) | 455.9 (255.7) | 425 (213.1) | 833.8 (355) | 6 |
| Part-time | 329.6 (70.5) | 402.1 (16.9) | 286.1 (66.5) | 834.6 (251.7) | 4 |
| Unemployed | 363.3 (269.6) | 467.5 (563.5) | 376.2 (257.4) | 1171 (1289) | 4 |
| Student | 215.1 (0) | 146.9 (0) | 256.6 (0) | 1207 (0) | 1 |
| **Marital Status** | | | | | |
| Single | 384.2 (235.8) | 486.6 (306.6) | 359.4 (225.6) | 1187 (881.4) | 7 |
| Cohabitation | - | - | - | - | 0 |
| Married | 363.2 (169.3) | 273.5 (186.6) | 367.6 (165.7) | 740.6 (337.3) | 8 |
| Divorced | - | - | - | - | 0 |
| **Residence** | | | | | |
| Parents | 325.6 (200.1) | 342.8 (247) | 303.8 (171.5) | 906.4 (349.7) | 6 |
| Alone | 318.5 (131.4) | 453.7 (119.3) | 318.5 (131.4) | 743.5 (372.6) | 6 |
| Roommate | - | - | - | - | 0 |
| Domestic Partner | 379.8 (186.9) | 513.2 (340.8) | 378.8 (188.2) | 706.3 (317.5) | 2 |

Table S3: Linear regression of Historical Leucine Levels and scores TRAQ, W-ADL, and WHOQOL-BREF, corrected for age.

| Mean Lifetime Leucine | | | |
|---|---|---|---|
| Measure | Regression slope | 95% CI | p-value |
| TRAQ - Total | -0.003 | -0.012 to 0.0055 | 0.45 |
| TRAQ - Medication Management | -0.0008 | -0.0034 to 0.0018 | 0.51 |
| TRAQ - Appointment Keeping | -0.0009 | -0.0041 to 0.0022 | 0.53 |
| TRAQ - Health Tracking | -0.001 | -0.004 to 0.0016 | 0.37 |
| TRAQ - Talking to Providers | -9.00E-05 | -0.001 to 0.00087 | 0.85 |
| W-ADL Total | 0.05 | -0.057 to 0.16 | 0.33 |
| WHOQOL - Environmental | 0.033 | -0.059 to 0.086 | 0.69 |
| WHOQOL - Physical | 0.045 | -0.022 to 0.11 | 0.17 |
| WHOQOL - Psychological | 0.024 | -0.063 to 0.11 | 0.56 |
| WHOQOL - Social | 0.019 | -0.041 to 0.08 | 0.5 |
| Mean Childhood Leucine | | | |
| Measure | Regression slope | 95% CI | p-value |
| TRAQ - Total | -0.0057 | -0.019 to 0.0081 | 0.34 |
| TRAQ - Medication Management | -0.0013 | -0.0057 to 0.0031 | 0.47 |
| TRAQ - Appointment Keeping | -0.0025 | -0.007 to 0.0021 | 0.23 |
| TRAQ - Health Tracking | -0.0017 | -0.0063 to 0.0029 | 0.38 |
| TRAQ - Talking to Providers | -0.00017 | -0.0021 to 0.0018 | 0.83 |
| W-ADL Total | 0.075 | -0.069 to 0.22 | 0.24 |
| WHOQOL - Environmental | 0.039 | -0.06 to 0.14 | 0.34 |
| WHOQOL - Physical | 0.034 | -0.022 to 0.09 | 0.16 |
| WHOQOL - Psychological | 0.031 | -0.064 to 0.13 | 0.42 |
| WHOQOL - Social | -0.0028 | -0.098 to 0.092 | 0.94 |
| Mean Adult Leucine | | | |
| Measure | Regression slope | 95% CI | p-value |
| TRAQ - Total | -0.00023 | -0.0095 to 0.009 | 0.96 |
| TRAQ - Medication Management | 4.51E-05 | -0.0027 to 0.0028 | 0.97 |
| TRAQ - Appointment Keeping | -3.69E-05 | -0.0035 to 0.0034 | 0.98 |
| TRAQ - Health Tracking | -0.00053 | -0.0036 to 0.0025 | 0.71 |

| | | | |
|---|---|---|---|
| TRAQ - Talking to Providers | 0.00029 | -0.0007 to 0.0013 | 0.53 |
| W-ADL Total | 0.043 | -0.071 to 0.16 | 0.43 |
| WHOQOL - Environmental | 0.008 | -0.07 to 0.086 | 0.83 |
| WHOQOL - Physical | 0.046 | -0.027 to 0.12 | 0.19 |
| WHOQOL - Psychological | 0.023 | -0.071 to 0.12 | 0.6 |
| WHOQOL - Social | 0.021 | -0.044 to 0.086 | 0.5 |
| Peak Leucine | | | |
| Measure | Regression slope | 95% CI | p-value |
| TRAQ - Total | -0.0027 | -0.0071 to 0.0017 | 0.2 |
| TRAQ - Medication Management | -0.00048 | -0.0019 to 0.0009 | 0.46 |
| TRAQ - Appointment Keeping | -0.0013 | -0.0029 to 0.00019 | 0.081 |
| TRAQ - Health Tracking | -0.00075 | -0.0022 to 0.00074 | 0.29 |
| TRAQ - Talking to Providers | -0.00014 | -0.00065 to 0.00036 | 0.55 |
| W-ADL Total | 0.03 | -0.027 to 0.087 | 0.27 |
| WHOQOL - Environmental | 0.02 | -0.017 to 0.058 | 0.26 |
| WHOQOL - Physical | 0.018 | -0.021 to 0.056 | 0.33 |
| WHOQOL - Psychological | 0.026 | -0.02 to 0.071 | 0.24 |
| WHOQOL - Social | -0.0032 | -0.037 to 0.0315 | 0.84 |

Figure S3

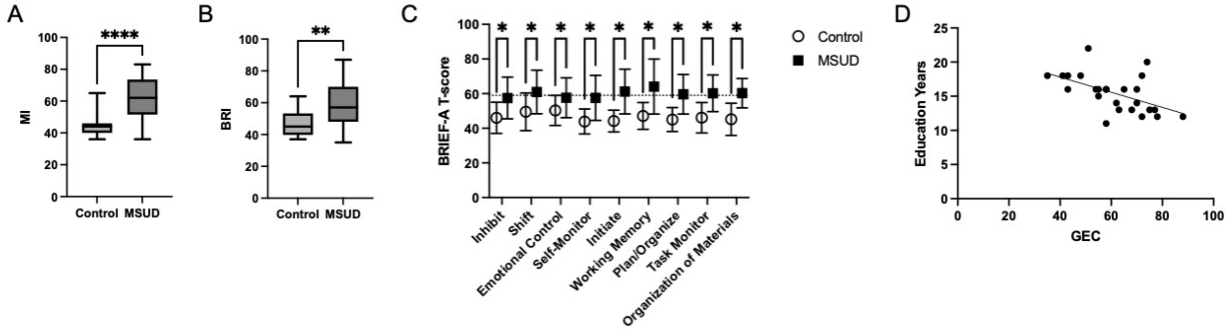


Fig S3. Subindices of the BRIEF-A questionnaire showed greater deficits in executive function in adults with MSUD and correlation between age-normalized scores and education. Adults with MSUD had significantly higher (A) Metacognitive Index (MI) T-scores (***$p<0.001$), (B) Behavioral Regulatory Index (BRI) T-scores (**$p=0.003$), and (C) subscales T-scores (*$p<0.01$) signifying greater executive function deficits. (D) Linear regression modeling, corrected for age, showed a correlation between lower T-scores (signifying better executive function) and more years of education completed (stats here).

*MI: Metacognitive Index; BRI: Behavioral Regulatory Index: GEC: General Executive Composite.*

Table S4: Linear regression of historic leucine levels and BRIEF-A T-scores, corrected for age.

| | | | |
|---|---|---|---|
| Mean Lifetime Leucine | | | |
| | Regression slope | 95% CI | p-value |
| GEC | -0.023 | -0.075 to 0.03 | 0.36 |
| MI | -0.038 | -0.092 to 0.016 | 0.15 |
| BRI | -0.0098 | -0.066 to 0.047 | 0.71 |
| Inhibit | -0.0047 | -0.058 to 0.048 | 0.85 |
| Shift | -0.019 | -0.07 to 0.032 | 0.44 |
| Emotional Control | 0.0075 | -0.043 to 0.058 | 0.75 |
| Self-Monitor | 0.017 | -0.034 to 0.068 | 0.47 |
| Initiate | -0.012 | -0.064 to 0.04 | 0.63 |
| Working Memory | -0.031 | -0.088 to 0.026 | 0.25 |
| Plan/Organize | -0.026 | -0.066 to 0.014 | 0.18 |
| Task Monitor | -0.013 | -0.055 to 0.028 | 0.49 |
| Organization of Materials | -0.036 | -0.073 to -0.00034 | 0.048 |
| Mean Childhood Leucine | | | |
| | Regression slope | 95% CI | p-value |
| GEC | -0.022 | -0.13 to 0.084 | 0.6 |
| MI | -0.023 | -0.12 to 0.071 | 0.53 |
| BRI | -0.016 | -0.12 to 0.089 | 0.7 |
| Inhibit | 0.00033 | -0.12 to 0.12 | 0.99 |
| Shift | -0.014 | -0.1 to 0.07 | 0.66 |
| Emotional Control | -0.0058 | -0.11 to 0.1 | 0.89 |
| Self-Monitor | 0.0015 | -0.14 to 0.14 | 0.98 |
| Initiate | 0.013 | -0.068 to 0.093 | 0.68 |
| Working Memory | -0.026 | -0.14 to 0.089 | 0.56 |
| Plan/Organize | -0.04 | -0.12 to 0.041 | 0.24 |
| Task Monitor | -0.044 | -0.11 to 0.019 | 0.13 |
| Organization of Materials | -0.014 | -0.093 to 0.064 | 0.64 |
| Mean Adult Leucine | | | |
| | Regression slope | 95% CI | p-value |
| GEC | -0.028 | -0.08115 to 0.02541 | 0.274 |

| | | | |
|---|---|---|---|
| MI | -0.043 | -0.097 to 0.012 | 0.11 |
| BRI | -0.015 | -0.073 to 0.042 | 0.57 |
| Inhibit | -0.019 | -0.072 to 0.035 | 0.46 |
| Shift | -0.025 | -0.076 to 0.026 | 0.31 |
| Emotional Control | 0.0005 | -0.052 to 0.053 | 0.98 |
| Self-Monitor | 0.0087 | -0.045 to 0.062 | 0.73 |
| Initiate | -0.014 | -0.067 to 0.039 | 0.57 |
| sWorking Memory | -0.037 | -0.094 to 0.021 | 0.19 |
| Plan/Organize | -0.025 | -0.067 to 0.016 | 0.2 |
| Task Monitor | -0.013 | -0.056 to 0.03 | 0.51 |
| Organization of Materials | -0.042 | -0.077 to -0.0069 | **0.023** |
| Peak Leucine | | | |
| | Regression slope | 95% CI | p-value |
| GEC | 0.0056 | -0.021 to 0.032 | 0.65 |
| MI | 0.0088 | -0.02 to 0.037 | 0.52 |
| BRI | 0.01 | -0.017 to 0.037 | 0.42 |
| Inhibit | 0.004 | -0.022 to 0.03 | 0.74 |
| Shift | 0.0032 | -0.022 to 0.029 | 0.79 |
| Emotional Control | 0.014 | -0.0092 to 0.037 | 0.21 |
| Self-Monitor | 0.018 | -0.0041 to 0.041 | 0.1 |
| Initiate | 0.001 | -0.025 to 0.027 | 0.93 |
| Working Memory | 0.0072 | -0.022 to 0.036 | 0.6 |
| Plan/Organize | 0.0019 | -0.019 to 0.023 | 0.85 |
| Task Monitor | -0.0064 | -0.027 to 0.014 | 0.5 |
| Organization of Materials | 0.0049 | -0.016 to 0.026 | 0.62 |

Figure S4

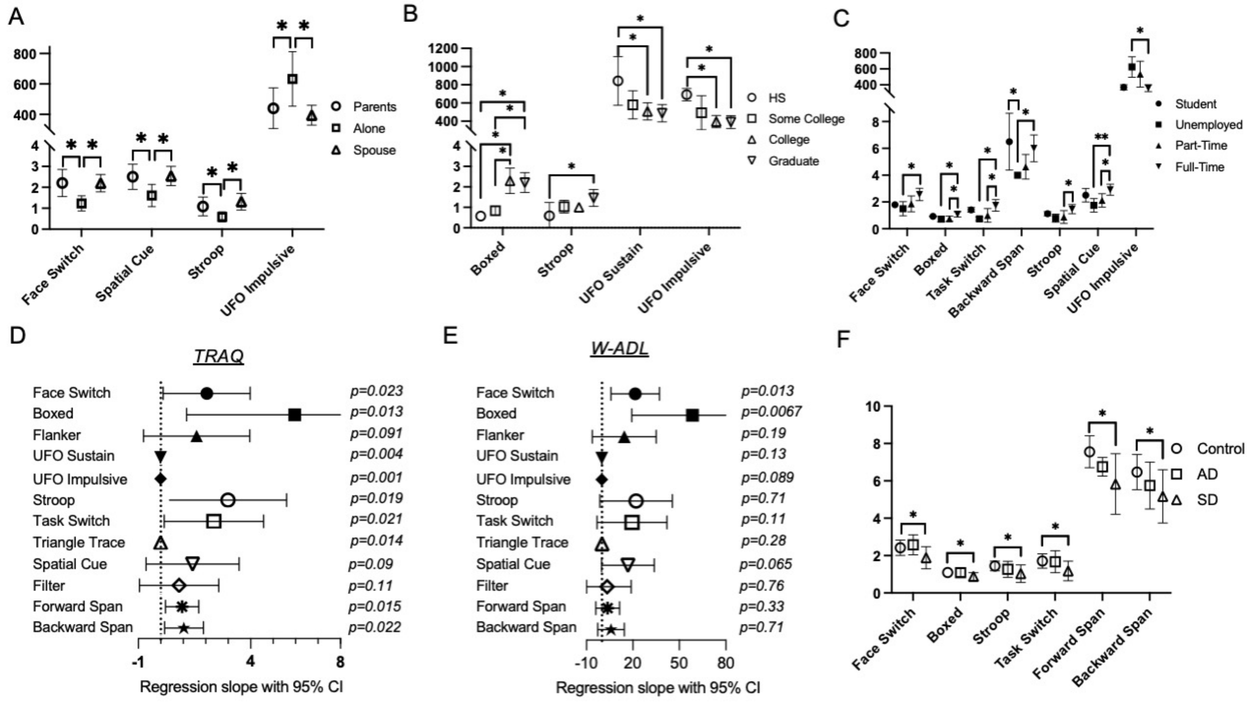


Fig S4: A subset of the ACE games correlate with psychosocial social outcomes and skills for independent living for adults with MSUD. (A) Adults with MSUD who live alone had significantly lower scores in 4 ACE games showing worse executive function: Face Switch (RCS), Spatial Cue (RCS), Stroop (RCS), UFO Impulsive (MRT) *$p<0.01$. All games not shown had no significant differences (B) Terminal degree correlated with higher scores (better executive function) on Boxed (RCS) for adults with college and graduate degrees compared to those with high school degrees. Adults with graduate degrees scored higher (better executive function) on Stroop (RCS) than adults who are high school graduates but did not attend college. Adults with graduate degrees had significantly lower MRTs (better executive function) in both attention games (UFO Sustain and UFO impulsive) than adults with terminal high school degrees. *$p<0.01$. All games not shown had no significant differences (C) ACE score correlates with employment in 7 games. Adults employed full-time had significantly higher RCS (better executive function) compared to unemployed individuals in Face Switch, Boxed, Task Switch, Spatial Cue *$p<0.01$. **$p<0.001$ Full-time employment correlated with significantly higher scores (better executive function) in Boxed, Task Switch, and Spatial Cue. Significantly greater MOS (better executive function) in Backward Span was seen in full-time employment versus unemployment and student versus unemployment *$p<0.01$. MRT in UFO Impulsive was significantly higher (worse executive function) for unemployed adults versus full-time employed adults *$p<0.01$. All games not shown had no significant differences (D-E) Forest plots are shown illustrating the regression slope with 95% confidence intervals and p-values for regression models adjusted for age between ACE game metric and total TRAQ scores (D) and scaled W-ADL scores (E). All p-values noted with an asterisk are significant beyond the Bonferroni-adjusted scale of 0.025. (F) Scores for 7 ACE games were significantly lower (worse executive function) for individuals diagnosed with MSUD symptomatically compared controls. This statistical difference compared to controls was absent for individuals diagnosed with MSUD prior to symptom appearance. Face Switch, Boxed, Stroop, and Task Switch all measured RCS. Forward Span and Backward Span measured MOS. **p<0.01.* All games not shown had no significant differences

*RCS: Rate Correct Score; MRT: Mean Reaction Time; MOS: Maximum Object Span; AD: Asymptomatic Diagnosis; SD: Symptomatic Diagnosis;*

Table S5: Linear regression of historic leucine levels and ACE scores, corrected for age.

| Mean Lifetime Leucine | | | |
|---|---|---|---|
| Game | Regression slope | 95% CI | p-value |
| Face switch | -0.00085 | -0.0051 to 0.0034 | 0.66 |
| Boxed | -0.0005 | -0.0015 to 0.00053 | 0.31 |
| Flanker | -0.00078 | -0.0032 to 0.0016 | 0.48 |
| UFO Sustain | 0.27 | -0.30 to 0.85 | 0.31 |
| UFO Impulsive | 0.52 | -0.46 to 1.49 | 0.26 |
| Stroop | 0.00074 | -0.0023 to 0.0038 | 0.6 |
| Task Switch | -0.00041 | -0.0045 to 0.0037 | 0.82 |
| TNT | 0.04 | -1.95 to 2.03 | 0.96 |
| Spatial Cue | -0.00058 | -0.0051 to 0.0039 | 0.78 |
| Filter | -0.0014 | -0.0073 to 0.0046 | 0.62 |
| Forward Span | -0.0022 | -0.0092 to 0.0048 | 0.5 |
| Backward Span | 0.0019 | -0.0029 to 0.0067 | 0.39 |
| Mean Childhood Leucine | | | |
| Game | Regression slope | 95% CI | p-value |
| Face switch | 0.00055 | -0.0056 to 0.0067 | 0.8 |
| Boxed | -0.00029 | -0.003 to 0.0024 | 0.75 |
| Flanker | 0.00037 | -0.0047 to 0.0055 | 0.83 |
| UFO Sustain | 0.3 | -0.7 to 1.30 | 0.41 |
| UFO Impulsive | 0.15 | -1.5 to 1.81 | 0.79 |
| Stroop | 0.00084 | -0.0023 to 0.004 | 0.46 |
| Task Switch | -0.00039 | -0.005 to 0.0042 | 0.75 |
| TNT | 0.29 | -2.72 to 3.31 | 0.78 |
| Spatial Cue | 0.00056 | -0.0049 to 0.006 | 0.76 |
| Filter | -0.002 | -0.01 to 0.007 | 0.52 |
| Forward Span | 0.0033 | -0.011 to 0.018 | 0.52 |
| Backward Span | -0.00046 | -0.011 to 0.018 | 0.9 |
| Mean Adult Leucine | | | |
| Game | Regression slope | 95% CI | p-value |
| Face switch | -0.00063 | -0.0051 to 0.0038 | 0.76 |

| | | | |
|---|---|---|---|
| Boxed | -0.00038 | -0.0014 to 0.00066 | 0.43 |
| Flanker | -0.00096 | -0.0033 to 0.0014 | 0.39 |
| UFO Sustain | 0.24 | -0.35 to 0.83 | 0.38 |
| UFO Impulsive | 0.24 | -0.87 to 1.34 | 0.64 |
| Stroop | 0.00047 | -0.0028 to 0.0037 | 0.75 |
| Task Switch | -0.00051 | -0.0047 to 0.0037 | 0.78 |
| TNT | -0.74 | -2.83 to 1.35 | 0.43 |
| Spatial Cue | -0.00062 | -0.0054 to 0.0041 | 0.77 |
| Filter | -0.00012 | -0.0065 to 0.0062 | 0.97 |
| Forward Span | -0.0016 | -0.0087 to 0.0054 | 0.62 |
| Backward Span | 0.0025 | -0.0021 to 0.0072 | 0.25 |
| Peak Leucine | | | |
| Game | Regression slope | 95% CI | p-value |
| Face switch | -0.00037 | -0.0018 to 0.0011 | 0.58 |
| Boxed | -0.00021 | -0.00077 to 0.00034 | 0.42 |
| Flanker | -0.00017 | -0.0016 to 0.0012 | 0.79 |
| UFO Sustain | -0.042 | -0.39 to 0.31 | 0.79 |
| UFO Impulsive | -0.068 | -0.46 to 0.32 | 0.67 |
| Stroop | -0.00014 | -0.0012 to 0.00095 | 0.77 |
| Task Switch | -0.00018 | -0.0017 to 0.0013 | 0.78 |
| TNT | 0.39 | -0.27 to 1.05 | 0.21 |
| Spatial Cue | -0.00024 | -0.0018 to 0.0013 | 0.74 |
| Filter | 0.00056 | -0.0015 to 0.0026 | 0.56 |
| Forward Span | 0.00079 | -0.003 to 0.0046 | 0.65 |
| Backward Span | -0.00033 | -0.003 to 0.0023 | 0.78 |

Figure S5

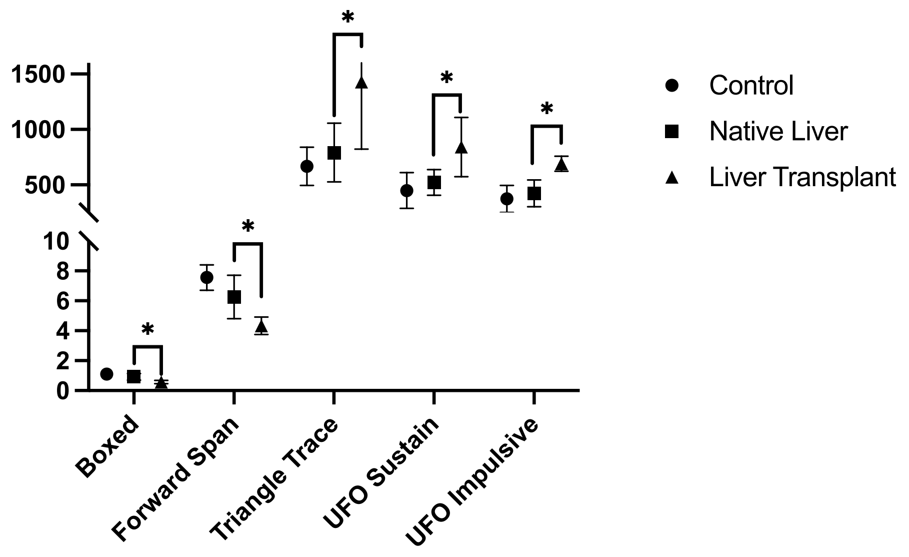


Figure S5: Subjects with history of liver transplantation showed significantly lower RCS scores (worse executive function) in Boxed and Forward Span compared to subjects with their native liver in Boxed and Forward span measured. Subjects with transplanted liver had significantly MRT higher scores (worse executive function) in Triangle Trace, UFO Sustained, and UFO Impulsive. All other games showed no significant difference between groups. **p<0.01.*
*RCS: Rate Correct Score; MRT: Mean Reaction Time*

Figure S6

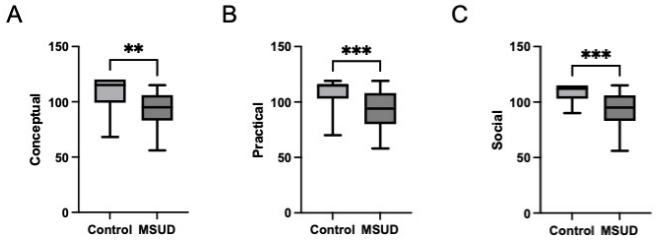


Figure S6: Affected subjects have worse adaptive function on ABAS-3 subdomains (A) Conceptual; (B) Practical; and (C) Social; compared to controls. ***p<0.005; ***p<0.0005*

Table S6: Linear regression of historic leucine levels and ABAS-3 scores, corrected for age.

| Mean Lifetime Leucine | | | |
|---|---|---|---|
| Index | Regression slope | 95% CI | p-value |
| GAC | 0.017 | -0.057-0.091 | 0.62 |
| Conceptual | -0.0017 | -0.078-0.074 | 0.96 |
| Practical | 0.031 | -0.045-0.11 | 0.39 |
| Social | 0.011 | -0.05-0.072 | 0.7 |
| Mean Childhood Leucine | | | |
| Index | Regression slope | 95% CI | p-value |
| GAC | -0.022 | --0.24-0.2 | 0.77 |
| Conceptual | -0.0056 | -0.24-0.23 | 0.94 |
| Practical | -0.009 | -0.24-0.22 | 0.91 |
| Social | -0.06 | -0.21-0.093 | 0.3 |
| Mean Adult Leucine | | | |
| Index | Regression slope | 95% CI | p-value |
| GAC | 0.031 | -0.041-0.01 | 0.36 |
| Conceptual | 0.012 | -0.063-0.088 | 0.72 |
| Practical | 0.043 | -0.03-0.12 | 0.22 |
| Social | 0.026 | -0.032-0.085 | 0.34 |
| Peak Leucine | | | |
| Index | Regression slope | 95% CI | p-value |
| GAC | -0.027 | -0.062-0.0079 | 0.11 |
| Conceptual | -0.02 | -0.058-0.018 | 0.28 |
| Practical | -0.031 | -0.067-0.0047 | 0.082 |
| Social | -0.017 | -0.047-0.014 | 0.24 |
